# Genetic Polymorphisms Associated with Insulin Resistance Risk in Normal BMI Indians

**DOI:** 10.1101/2024.08.22.24311638

**Authors:** Sabitha Thummala, Junaid Ahmed Khan Ghori, Katherine Saikia, Sarah Fathima, Nithya Kruthi, AR Balamurali, Rahul Ranganathan

## Abstract

**Background:** Insulin resistance (IR) contributes significantly to the onset of metabolic disorders, such as type 2 diabetes mellitus and cardiovascular diseases. Identifying genetic markers associated with IR can offer insights into its mechanisms and potential therapeutic targets.

**Objective:** This study investigated the association between four single nucleotide polymorphisms (SNPs) and insulin resistance among 191 individuals in the Indian population.

**Methods:** A literature review identified four SNPs linked to IR. Participants were divided into groups based on insulin resistance and sensitivity, determined by the Homeostasis Model Assessment for Insulin Resistance (HOMA2-IR). DNA was extracted for genotyping using Illumina Infinium Global Screening Array (GSA) V3. Case-control analysis assessed SNP-genotype associations with insulin resistance and other clinical parameters.

**Results:** Among 191 participants, 57 were insulin-resistant and 134 were insulin-sensitive. Significant associations (P < 0.05) were found between selected SNPs and IR. SNP rs920590 showed the strongest association, with the T allele associated with increased IR risk (odds ratio = 4.01, 95% CI 1.55-10.34; p < 0.0014). Additionally, serum LDL cholesterol, serum triglycerides, HbA1c, Insulin fasting and fat mass show significant differences in cases and controls.

**Conclusion:** This study validates genetic markers linked to insulin resistance (IR) in the Indian population and elucidates their roles in IR pathogenesis. Understanding these markers can inform personalised therapeutic strategies for metabolic disorders.

## INTRODUCTION

Diabetes Mellitus (DM) has a rich historical record dating back approximately 3000 years to Egyptian manuscripts^1^, making it one of humanity’s oldest known diseases. It encompasses a spectrum of metabolic disorders involving lipid, protein, and carbohydrate metabolism and is characterised by persistent hyperglycemia^2^. In 2019, noncommunicable diseases (NCDs) accounted for 74% of global deaths, with diabetes alone contributing to 1.6 million fatalities^3, 4^. This global burden is particularly seen in developing countries like India, where an estimated 77 million individuals were affected by diabetes in 2019^4^. Notably, individuals with Diabetes Mellitus face a significantly heightened risk of developing Metabolic Syndrome (Met S)^5^.

Traditionally, Body mass index (BMI) has been considered a key marker of metabolic syndrome. However, recent research suggests the involvement of additional factors^6, 7^. Higher BMI does not consistently correlate with a higher frequency of metabolic syndrome among Asian patients^8^, prompting a reevaluation of conventional metrics. Focusing on body composition, particularly muscle and fat mass, provides a more nuanced understanding of metabolic health^9^. Previous epidemiological research has explored the relationship between fat mass and insulin resistance^10^. Anthropometric measures such as waist circumference and waist-to-hip ratio have emerged as better predictors of obesity-associated type 2 diabetes risk than BMI alone^11, 12^. However, the influence of body fat percentage (BF%) on insulin resistance remains underexplored, especially among individuals with normal BMI^13, 14^. Recent genetic research has identified correlations between specific genetic loci and metabolic traits, shedding light on the genetic underpinnings of metabolic health disparities^15^. Building upon this foundation, our study aims to delve into the potential genetic influences on insulin resistance within the Indian population, particularly among individuals with normal BMI. Through a comprehensive literature survey, we have identified a set of genes and SNPs linked with insulin resistance (IR). We seek to validate these findings within our population and ascertain their relevance in the Indian context.

Given the emerging evidence suggesting genetic correlations with metabolic traits, our study seeks to explore the potential impact of genetic polymorphisms in four specific genes (*INTS10, LINC01427–LINC00261, SENP2*, and *SLC22A11*) on insulin resistance among individuals with normal BMI in the Indian population^16^. By investigating the association between these genetic variants and insulin resistance risk, we aim to enhance our understanding of the genetic underpinnings of metabolic health indicators. Recent genetic research has shown correlations between particular loci and percentage of body fat, suggesting protective effects against glycaemic and lipid outcomes^17^. Lowered total adiposity is typically associated with increased genetic susceptibility to insulin resistance, indicating that poor fat storage may be detrimental to metabolism. We will concurrently examine four SNPs within these genes: rs920590 in *INTS10*, rs7274134 in *LINC01427 - LINC00261*, rs6762208 in *SENP2*, and rs2078267 in *SLC22A11*.

## MATERIALS AND METHODS

### 1. Study Participants

The research enrolled 191 participants (90 men and 101 women) of Indian ethnicity residing in India, aged 18-65 years, between January 2022 and December 2023. HOMA2-IR was used to categorise participants into cases and controls. Individuals with HOMA2-IR > 2 were classified as cases (57 participants), and those with HOMA2-IR < 2 (134 participants) were categorised as controls. The HOMA2-IR was calculated using the online tool provided by the Medical Science Division of The University of Oxford^18^. An electronic registration form was used to gather the demographic data of the participants, including their self-reported weight (kg) and height (cm). The formula for calculating body mass index (BMI) was weight (kg)/height (m^2)^19^. For adult males, the formula for Body Fat Percentage (BFP) was 1.20 x BMI + 0.23 x Age - 16.2, for an adult female, the formulas for Body Fat Percentage (BFP) were 1.20 x BMI + 0.23 x Age - 5.420^20^ and Fat Mass was BFP x Weight x 0.0120^20^. To ensure participant safety and ethical consideration, the study included only individuals free from: cancer history, cardiovascular or renal failure, mental illnesses, and pregnancy or lactation. Every participant gave written, informed consent in accordance with the guidelines in the 2008 updated Declaration of Helsinki^21^. The study protocol received approval from the Answer Genomics Ethical Review Committee. Figure 1 provides a detailed graphical representation of the study’s structure and key components, outlining the research methodology, participant selection, and analysis framework.

**Figure 1:**
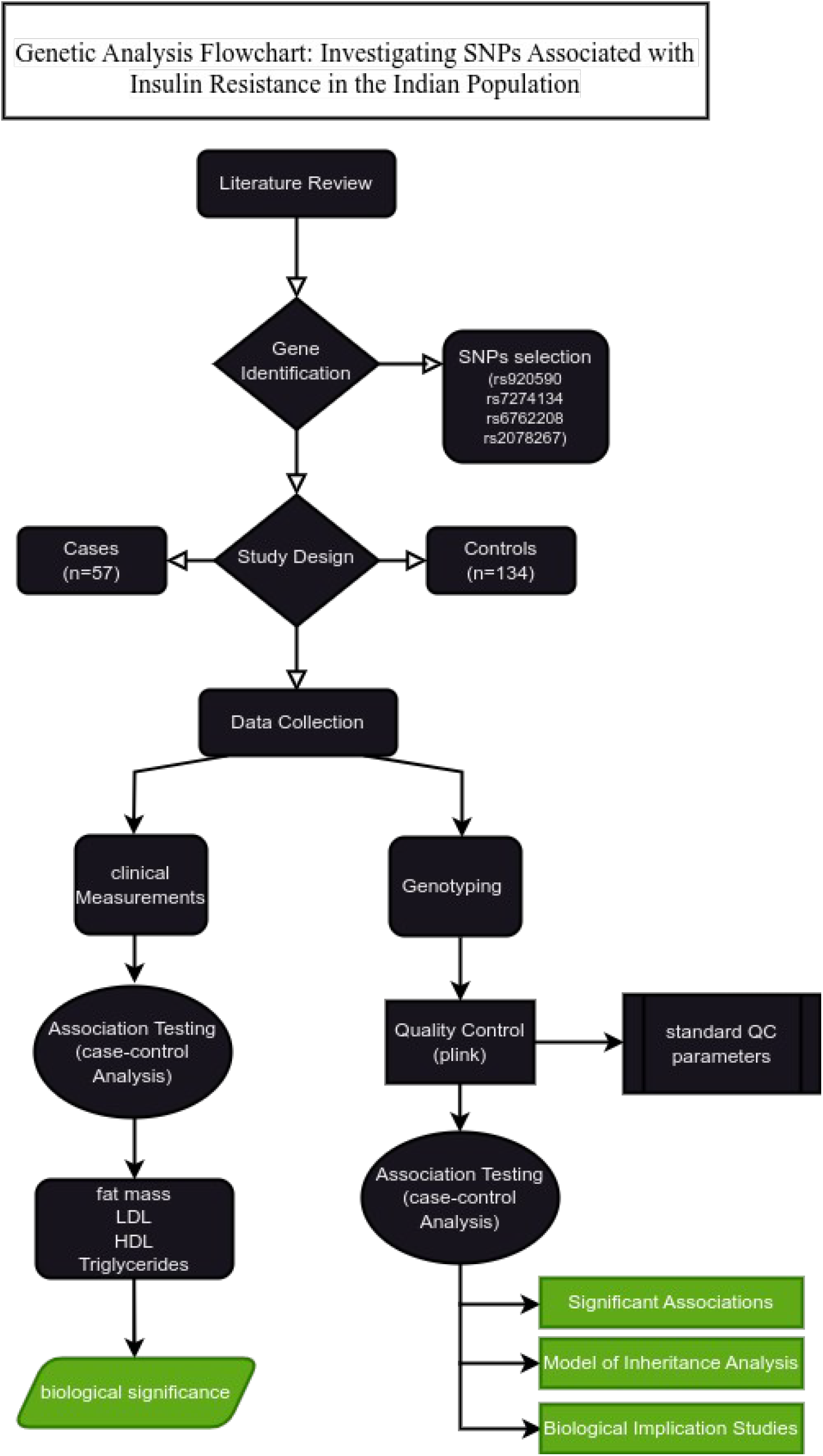
Study Flowchart.

### 2. Laboratory Measurements

Following a 12-hour fast, participants’ blood was drawn for analysis of important metabolic markers, including, high-density lipoprotein cholesterol (LDL-C), total cholesterol (TC), triglycerides (TG), fasting plasma glucose (FPG), and low-density lipoprotein cholesterol (LDL-C),. The assessments were conducted using Beckman DxC 700 AU for all markers except HbA1c, which was analyzed with Tosoh G-8, and fasting insulin levels measured by Beckman UniCel DxI 800, adhering to the protocols provided by the manufacturers. The healthy reference ranges for these markers were established as follows: FPG (70-100 mg/dl), TC (0-100 mg/dl), TG (0-150 mg/dl), HDL-C (40-60 mg/dl), and LDL-C (0-100 mg/dl). To determine insulin sensitivity, the Homeostasis Model Assessment of Insulin Resistance (HOMA2-IR) was applied, calculated from fasting plasma glucose (in mmol/l) and fasting insulin (in mU/l) using the formula: fasting glucose times fasting insulin/22.5^10^. Participants with a HOMA2-IR score above 2 were categorized as insulin resistant, while those with scores below 2 were considered insulin sensitive. The clinical characteristics of the study subjects and the distribution of these groupings are detailed in Table 2.

**Table 1:**
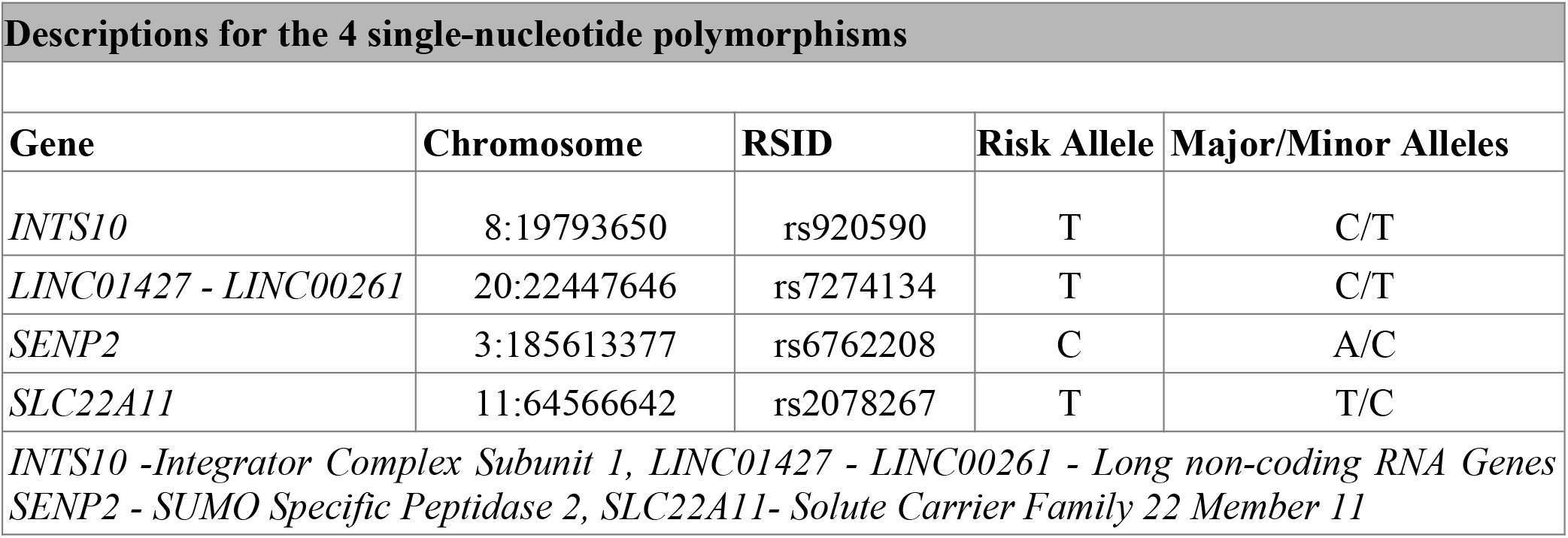
Overview of the four single-nucleotide polymorphisms.

**Table 2:** Clinical Profiles and Glucose and Lipid Metabolism Parameters of Participants in the Current Study.

|  |  |  |  |  |
| --- | --- | --- | --- | --- |
| Parameter | Cases Median (Q1-Q3) | Controls Median (Q1-Q3) | U Statistic | P Value |
| Age<sup>a</sup> | 30.0 (27.0-37.0) | 31.0 (27.0-38.0) | 3758.5 | 0.86 |
| Gender | 30.14136,26.85864 | 70.85864,63.14136 |  | 0.667 |
| Body Mass Index<sup>a</sup> | 23.05 (21.89-24.09) | 22.235 (20.84-23.61) | 4656.5 | 0.01 |
| Body Fat Percentage<sup>a</sup> | 26.11 (19.47-29.01) | 23.02 (17.9-27.52) | 4409.5 | 0.09 |
| Fat Mass<sup>a</sup> | 15.85 (13.59-17.79) | 13.62(11.78-16.04) | 4761.5 | 0.007 |
| HBA1C<sup>a</sup> | 5.4 (5.3-5.5) | 5.3 (5.1-5.5) | 4695.5 | 0.01 |
| Insulin Fasting<sup>a</sup> | 11.02 (9.67-13.88) | 5.31(3.9-6.92) | 7143 | 1.96E-21 |
| Glucose Fasting<sup>a</sup> | 92.6 (86.9-96.2) | 87.05 (82.42-92.27) | 4937.5 | 0.001 |
| Total Cholesterol<sup>a</sup> | 186.0 (166.0-227.0) | 185.0 (161.25-211.0) | 4146 | 0.35 |
| Serum LDL Cholestrol<sup>a</sup> | 46.5 (42.9-52.5) | 49.5 (44.03-54.68) | 3383.0 | 2.128e-01 |

| Parameter | Cases Median (Q1-Q3) | Controls Median (Q1-Q3) | U Statistic | P Value |
| --- | --- | --- | --- | --- |
| Age<sup>a</sup> | 30.0 (27.0-37.0) | 31.0 (27.0-38.0) | 3758.5 | 0.86 |
| Gender | 30.14136,26.85864 | 70.85864,63.14136 |  | 0.667 |
| Body Mass Index<sup>a</sup> | 23.05 (21.89-24.09) | 22.235 (20.84-23.61) | 4656.5 | 0.01 |
| Body Fat Percentage<sup>a</sup> | 26.11 (19.47-29.01) | 23.02 (17.9-27.52) | 4409.5 | 0.09 |
| Fat Mass<sup>a</sup> | 15.85 (13.59-17.79) | 13.62(11.78-16.04) | 4761.5 | 0.007 |
| HbA1C<sup>a</sup> | 5.4 (5.3-5.5) | 5.3 (5.1-5.5) | 4695.5 | 0.01 |
| Insulin Fasting<sup>a</sup> | 11.02 (9.67-13.88) | 5.31(3.9-6.92) | 7143 | 1.96E-21 |
| Glucose Fasting<sup>a</sup> | 92.6 (86.9-96.2) | 87.05 (82.42-92.27) | 4937.5 | 0.001 |
| Serum Triglycerides<sup>a</sup> | 102.0 (85.0-127.0) | 88.0 (70.0-126.0) | 4631 | 0.02 |
| Serum HDL Cholesterol<sup>a</sup> | 46.5 (42.9-52.5) | 49.5 (44.025-54.67) | 3383 | 0.21 |

<sup>a</sup>Mann-Whitney test; $\chi^2$ test. Data are presented as the median (Q1-Q3) for age, BMI, Fat Mass,HbA1c, total cholesterol, triglycerides, HDL, LDL, Glucose fasting, Insulin Fasting. Data are presented as n for gender. P<0.05 was considered to indicate a statistically significant difference.

### 3. Genotyping and Single-nucleotide polymorphism(SNP) selection

This study investigated potential genetic influences on insulin resistance by analyzing participants’ DNA. The extraction of DNA was conducted on blood specimens utilizing the Qiagen blood extraction kit, acclaimed for its capability to isolate genomic DNA of superior quality from blood samples. The process of genotyping SNPs was executed with the aid of the Illumina Infinium Global Screening Array (GSA) V3 platform, recognized for its extensive representation of genetic variance throughout the genome^22^. The Illumina iScan system was used for genotyping, while Genome Studio V2 software was used for interpreting the raw intensity data, quality control, and data export^23^.

The study focused on four genes (*INTS10, LINC01427 - LINC00261, SENP2* and *SLC22A11*) involved in metabolism and potentially influencing insulin resistance, as indicated in Table 1. The selection of SNPs was based on a comprehensive literature review of scientific literature identifying genetic markers associated with metabolic traits.

### 4. Statistical analysis

The evaluation of statistical differences commenced by examining the variations in age, glucose, and lipid metrics such as LDL cholesterol, HDL cholesterol, total cholesterol, triglycerides, and HbA1c between two groups. Given the data’s non-parametric characteristics, the Mann-Whitney U test (utilising the Scipy library) was utilised for these analyses^17^. Gender distribution across the groups was evaluated using the Chi-square test, with detailed results presented in Table 2.

Utilising PLINK software^24^ for quality control, the study filtered genotype data by removing samples with low genotyping rates, excluding SNPs with significant missing data, and eliminating rare variants for their minimal statistical power. Heterozygosity checks were also conducted to ensure accurate genotype distribution. Subsequently, a study was carried out using SNPstats software^25^ to ascertain the correlation between the described four SNPs and insulin resistance. The analysis yielded odds ratios (ORs) together with their 95% confidence intervals (CIs). Table 3 summarises the results, which show significant SNP markers under different genetic inheritance models.

**Table 3:**
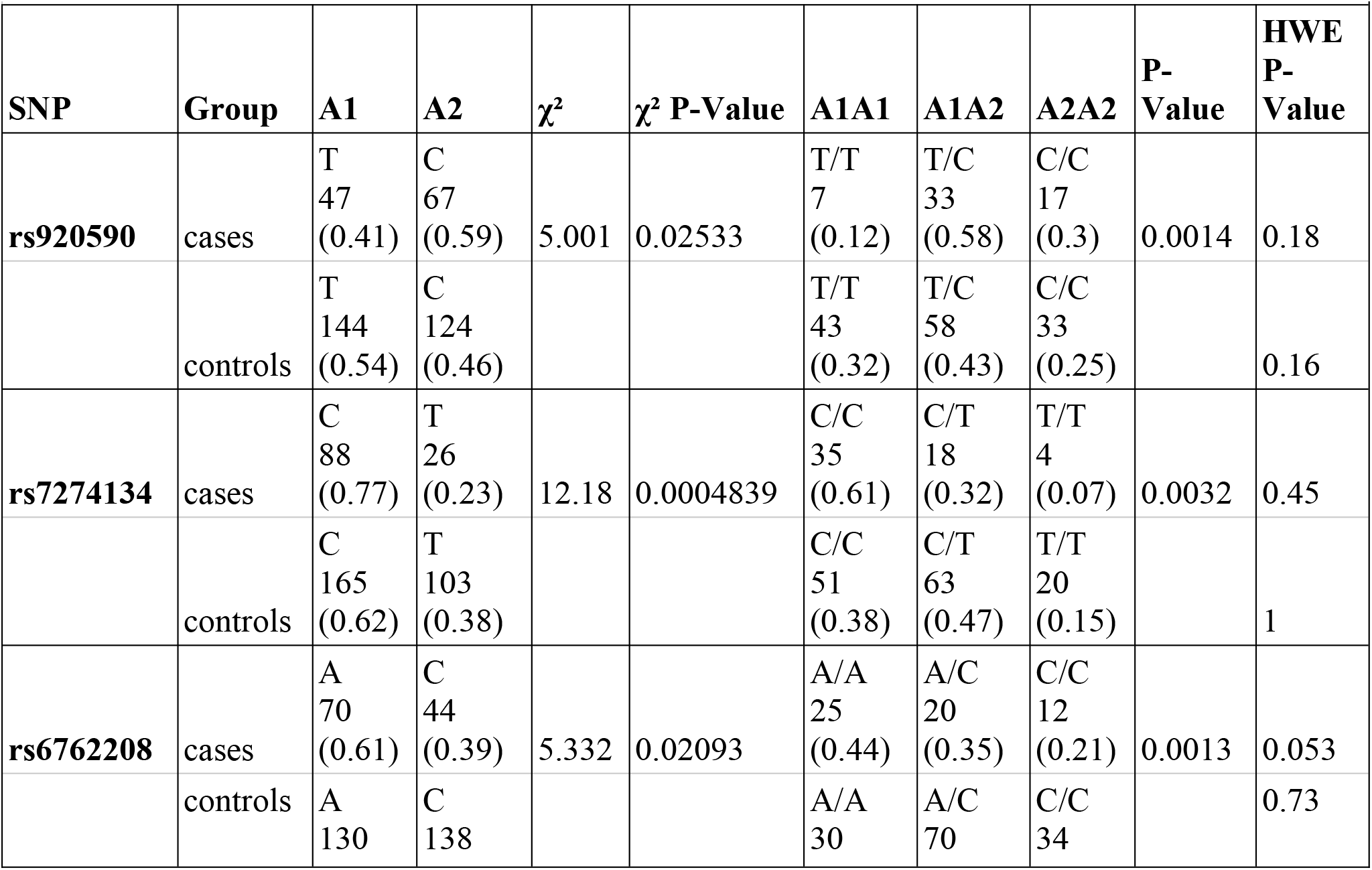
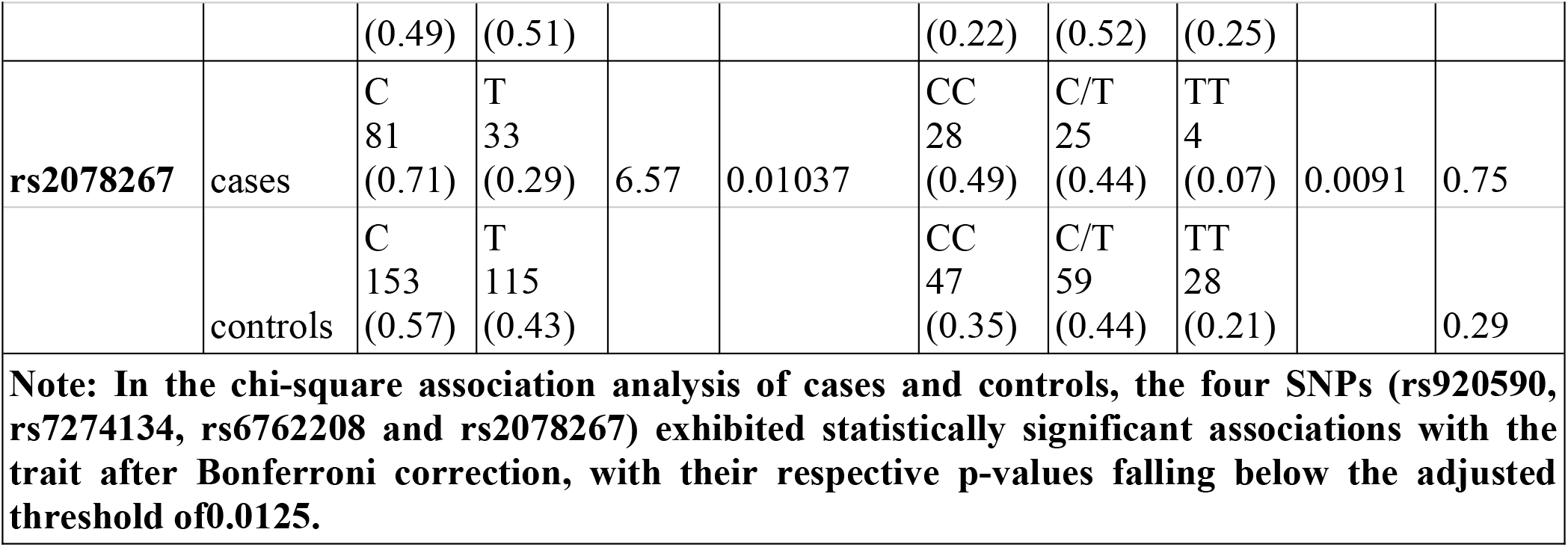
Comparison of allelic and genotypic distribution of four SNPs (rs920590, rs7274134, rs6762208 and rs2078267) between the two groups HOMA2 IR >2 and HOMA2 IR <2.

In the control group, Hardy-Weinberg equilibrium was maintained with a significance level set at 0.05. The assessment of risk was conducted through the calculation of odds ratios (OR) along with their 95% confidence intervals (95% CI). The study also evaluated the linkage disequilibrium (LD) between the SNPs, with the LD coefficient D being determined via the use of Snpstats software ^25^. Additionally, power analyses were conducted based on power and sample size calculations^26^. Each SNP underwent a Bonferroni adjustment was carried out for each SNP to rule out the possibility of a type I error brought on by multiple tests.

## RESULTS

### Subject characteristics

In this study, all 191 participants were genotyped, with their clinical features and lipid metabolism indicators detailed in Table 2. The investigation revealed no meaningful statistical disparities in age or gender between the case and control groups, nor among males and females within those groups (p>0.05). Yet, notable distinctions were observed in BMI (0.01), Fat Mass (0.007), HBA1C (0.01), Glucose Fasting (0.001), and Serum Triglycerides (0.02), with the exceptions of BFP, Total Cholesterol, and Serum HDL Cholesterol when comparing cases to controls.

### Association of the 4 SNPs with IR

In this study, the relationship between IR and four SNPs in the Indian population’s normal weight individuals was examined. To ensure the study had sufficient statistical power, a sample size calculation was performed. This calculation aimed to detect significant associations, considering a significance level (alpha) of 0.05.

The frequency of genotypes for all four SNPs among both cases and controls was evaluated to examine their compliance with Hardy-Weinberg equilibrium (HWE) through a Fisher exact test. None of the SNPs demonstrated a significant deviation from HWE (p > 0.05).suggesting the absence of genotyping errors and population stratification, the results are mentioned in Table 3.

We performed chi-square tests to evaluate the association between genotypes and allele frequencies of each SNP with phenotype (IR). After Bonferroni correction for multiple testing (p < 0.0125), all four SNPs displayed significant associations with IR:

Chi-squared analyses were performed to investigate the relationship between the genotypic and allelic variations of four SNPs (rs920590, rs7274134, rs6762208, and rs2078267) within two groups distinguished by their HOMA2 IR levels (>2 and <2).The obtained p-values indicate significant associations between SNP genotypes and insulin resistance status.The allelic frequency difference between the cases and control were also found to be significant (p< 0.05)

- For rs920590 (*INTS10*), individuals carrying the T/T genotype had an elevated risk of IR compared to those with C/C-C/T genotypes [Odds Ratio (OR) = 4.01; 95% Confidence Interval (CI): 1.55-10.34; p < 0.0014] under the recessive model.
- rs7274134 (*LINC01427 - LINC00261*), individuals carrying the C/T-T/T genotypes had an elevated risk of IR [Odds Ratio (OR) = 2.60, 95% CI: 1.37-4.96, p < 0.0032] according to the dominant model.
- rs6762208 (*SENP2*), individuals carrying the A/C-C/C genotypes were associated with an increased risk of IR [Odds Ratio (OR) = 3.11, 95% CI: 1.55-6.24, p < 0.0013] under the dominant model.
- rs2078267 (*SLC22A11*), individuals carrying the T/T genotype showed a higher risk of IR [Odds Ratio (OR) = 3.72, 95% CI: 1.22-11.28, p < 0.0091] under the recessive model.

## MODEL OF INHERITANCE ANALYSIS

In order to comprehend the connection between genotype and insulin resistance (IR) risk in the Indian population, genetic inheritance patterns for the chosen SNPs were analyzed. Tables 4 through 7 outline the various inheritance models examined, including codominant, dominant, recessive, overdominant, and log-additive models, for SNPs rs920590 (*INTS10*), rs7274134 (*LINC01427 - LINC00261*), rs6762208 (*SENP2*), and rs2078267 (*SLC22A11*).

**Table 4.**
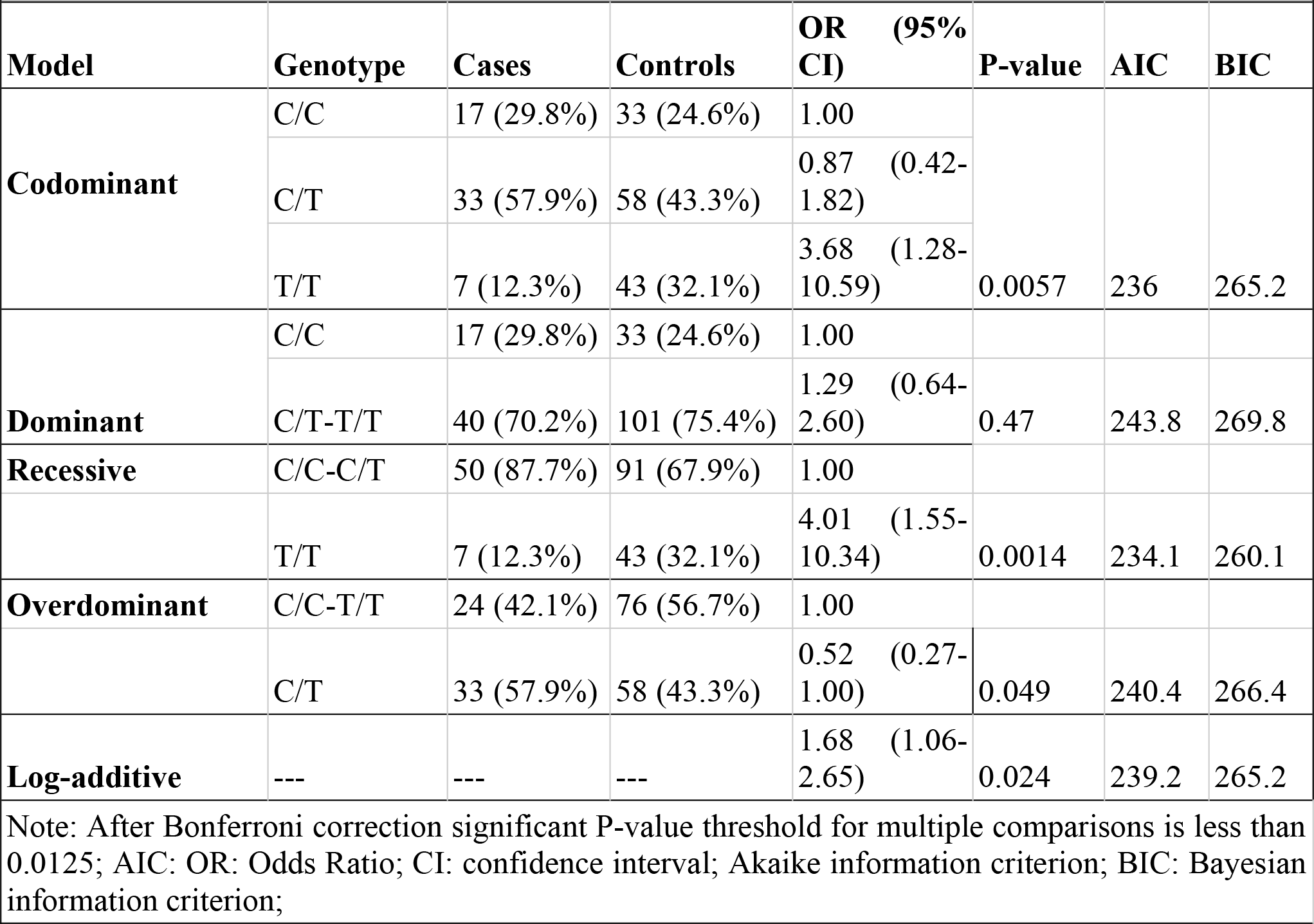
Inheritance models analysis of the SNP rs920590 (INTS10) between the Cases and Controls rs920590 association with response status (n=191, adjusted by gender+age.cat.

| Model | Genotype | Cases | Controls | OR (95% CI) | P-value | AIC | BIC |
| --- | --- | --- | --- | --- | --- | --- | --- |
| **Codominant** | C/C | 17 (29.8%) | 33 (24.6%) | 1.00 | 0.0057 | 236 | 265.2 |
|  | C/T | 33 (57.9%) | 58 (43.3%) | 0.87 (0.42-1.82) |  |  |  |
|  | T/T | 7 (12.3%) | 43 (32.1%) | 3.68 (1.28-10.59) |  |  |  |
| **Dominant** | C/C | 17 (29.8%) | 33 (24.6%) | 1.00 | 0.47 | 243.8 | 269.8 |
|  | C/T-T/T | 40 (70.2%) | 101 (75.4%) | 1.29 (0.64-2.60) |  |  |  |
| **Recessive** | C/C-C/T | 50 (87.7%) | 91 (67.9%) | 1.00 | 0.0014 | 234.1 | 260.1 |
|  | T/T | 7 (12.3%) | 43 (32.1%) | 4.01 (1.55-10.34) |  |  |  |
| **Overdominant** | C/C-T/T | 24 (42.1%) | 76 (56.7%) | 1.00 | 0.049 | 240.4 | 266.4 |
|  | C/T | 33 (57.9%) | 58 (43.3%) | 0.52 (0.27-1.00) |  |  |  |
| **Log-additive** | --- | --- | --- | 1.68 (1.06-2.65) | 0.024 | 239.2 | 265.2 |

Note: After Bonferroni correction significant P-value threshold for multiple comparisons is less than 0.0125; AIC: OR: Odds Ratio; CI: confidence interval; Akaike information criterion; BIC: Bayesian information criterion;

**Table 5.** Inheritance models analysis of the SNP rs7274134 (*LINC01427 - LINC00261*) between the Cases and Controls rs7274134 association with response status (n=191, adjusted by gender+age.cat)

| *Model* | *Genotype* | *Cases* | *Controls* | *OR (95% CI)* | *P-value* | *AIC* | *BIC* |
| --- | --- | --- | --- | --- | --- | --- | --- |
|  | C/C | 35 (61.4%) | 51 (38.1%) | 1.00 |  |  |  |
| **Codominant** | C/T | 18 (31.6%) | 63 (47%) | 2.43 (1.22-4.83) | 0.011 | 237.3 | 266.6 |
|  | T/T | 4 (7%) | 20 (14.9%) | 3.40 (1.03-11.24) |  |  |  |
| **Dominant** | C/C | 35 (61.4%) | 51 (38.1%) | 1.00 |  |  |  |
|  | C/T-T/T | 22 (38.6%) | 83 (61.9%) | 2.60 (1.37-4.96) | 0.0032 | 235.6 | 261.6 |
| **Recessive** | C/C-C/T | 53 (93%) | 114 (85.1%) | 1.00 |  |  |  |
|  | T/T | 4 (7%) | 20 (14.9%) | 2.35 (0.73-7.56) | 0.13 | 241.9 | 268 |
| **Overdominant** | C/C-T/T | 39 (68.4%) | 71 (53%) | 1.00 |  |  |  |
|  | C/T | 18 (31.6%) | 63 (47%) | 2.00 (1.03-3.91) | 0.038 | 240 | 266 |
| **Log-additive** | — | — | — | 2.08 (1.25-3.48) | 0.0034 | 235.7 | 261.8 |

**Table 6.** Inheritance models analysis of the SNP rs6762208 (*SENP2*) between the Cases and Controls rs6762208 association with response status (n=191, adjusted by gender+age.cat)

| *Model* | *Genotype* | *Cases* | *Controls* | *OR (95% CI)* | *P-value* | *AIC* | *BIC* |
| --- | --- | --- | --- | --- | --- | --- | --- |
|  | A/A | 25 (43.9%) | 30 (22.4%) | 1.00 |  |  |  |
| **Codominant** | A/C | 20 (35.1%) | 70 (52.2%) | 3.29 (1.55-7.00) | **0.0054** | 235.9 | 265.1 |
|  | C/C | 12 (21.1%) | 34 (25.4%) | 2.80 (1.16-6.77) |  |  |  |
| **Dominant** | A/A | 25 (43.9%) | 30 (22.4%) | 1.00 |  |  |  |
|  | A/C-C/C | 32 (56.1%) | 104 (77.6%) | 3.11 (1.55-6.24) | 0.0013 | 234 | 260 |
| **Recessive** | A/A-A/C | 45 (79%) | 100 (74.6%) | 1.00 |  |  |  |
|  | C/C | 12 (21.1%) | 34 (25.4%) | 1.35 (0.63-2.90) | 0.43 | 243.7 | 269.7 |
| **Overdominant** | A/A-C/C | 37 (64.9%) | 64 (47.8%) | 1.00 |  |  |  |
|  | A/C | 20 (35.1%) | 70 (52.2%) | 2.08 (1.08-3.97) | 0.025 | 239.3 | 265.3 |
| **Log-additive** | — | — | — | 1.79 (1.13-2.84) | 0.012 | 237.9 | 264 |

**Table 7.** Inheritance models analysis of the SNP rs2078267 (*SLC22A11*) between the Cases and Controls rs2078267 association with response status (n=191, adjusted by gender+age.cat)

| *Model* | *Genotype* | *Cases* | *Controls* | *OR (95% CI)* | *P-value* | *AIC* | *BIC* |
| --- | --- | --- | --- | --- | --- | --- | --- |
|  | C/C | 28 (49.1%) | 47 (35.1%) | 1.00 |  |  |  |
| ***Codominant*** | C/T | 25 (43.9%) | 59 (44%) | 1.38 (0.70-2.72) | **0.022** | 238.6 | 267.9 |
|  | T/T | 4 (7%) | 28 (20.9%) | 4.36 (1.37-13.92) |  |  |  |
| ***Dominant*** | C/C | 28 (49.1%) | 47 (35.1%) | 1.00 |  |  |  |
|  | C/T-T/T | 29 (50.9%) | 87 (64.9%) | 1.80 (0.94-3.41) | 0.074 | 241.1 | 267.1 |
| ***Recessive*** | C/C-C/T | 53 (93%) | 106 (79.1%) | 1.00 |  |  |  |
|  | T/T | 4 (7%) | 28 (20.9%) | 3.72 (1.22-11.28) | 0.0091 | 237.5 | 263.5 |
| ***Overdominant*** | C/C-T/T | 32 (56.1%) | 75 (56%) | 1.00 |  |  |  |
|  | C/T | 25 (43.9%) | 59 (44%) | 0.97 (0.51-1.84) | 0.93 | 244.3 | 270.3 |
| ***Log-additive*** | — | — | — | 1.81 (1.13-2.90) | 0.011 | 237.9 | 263.9 |

The determination of the optimal inheritance model for each SNP was based on identifying the model with the lowest Akaike Information Criterion (AIC) and Bayesian Information Criterion (BIC) values. For rs7274134 in *LINC01427 - LINC00261* and rs6762208 in *SENP2*, the dominant model exhibited the lowest AIC, indicating its suitability for describing the inheritance pattern. Specifically, the risk genotype for IR associated with rs7274134 was (C/T-T/T), with a significant association (P=0.0032, OR=2.60; 95% CI: 1.37-4.96). Similarly, the risk genotype for IR linked to rs6762208 was (A/C-C/C), showing a strong correlation (P=0.0013, OR=3.11; 95% CI: 1.55-6.24).

Conversely, for rs920590 in *INTS10* and rs2078267 in *SLC22A11*, the recessive model demonstrated the lowest AIC. In the case of rs920590, the risk genotype (T/T) was identified against the C/C-C/T genotypes for IR, with significant results (P=0.0014; OR=4.01; 95% CI: 1.55-10.34). Similarly, for rs2078267, the T/T genotype was marked as a risk factor compared to the C/C-C/T genotypes, with statistical significance (P=0.0091; OR=3.72; 95% CI: 1.22-11.28).

These findings support the role of these genetic markers in predisposing individuals to insulin resistance. Understanding the underlying mechanisms associated with these genetic variants could provide valuable insights into the development of personalised treatment approaches for metabolic disorders in the Indian population.

## DISCUSSION

This study investigated the association between specific genetic markers and insulin resistance (IR) among individuals with normal BMI in the Indian population, validating the role of SNPs (rs920590 in *INTS10*, rs7274134 in *LINC01427* - *LINC00261*, rs6762208 in *SENP2*, and rs2078267 in *SLC22A11*) in predisposing individuals to IR. These findings underscore the importance of genetic factors in metabolic dysregulation, validating previous research linking these genetic variants to metabolic traits.

### INTS10

Protein coding gene *INTS10* (Integrator Complex Subunit 10) is a part of the Integrator (INT) complex, which is a complex engaged in the transcription pathway leading to gene expression^27^. In a diabetic retinopathy study done on mice, it was found that the transcription of *INTS10* in the inner retinal cells of diabetes induced mice was upregulated early, suggesting that *INST10* contributes to diabetic retina due to high insulin resistance^28^. In another GWAS study carried out on the African population, a polymorphism was reported to be associated with BMI that lies between *INTS10* and *LPL* genes, which the study suggests is a BMI loci^29^. *LPL* gene has Lipoprotein lipase activity that is crucial in amassing the triglycerides from the blood and storing them in the adipocytes, which is seen more in women than in men^30^. Serum LPL mass is linked to metabolic syndrome, coronary heart disease, and ultimately insulin sensitivity, according to a study by Onat et al.^31^.

### LINC01427 - LINC00261

With more than 200 nucleotides, long non-coding RNAs (lncRNAs) are a type of significant regulatory RNAs that are involved in a variety of cellular functions in both healthy and pathological conditions.^32, 33^. They affect translation processes, RNA splicing, stability, chromatin structure and function, and gene transcription. Additionally, they play a role in the formation and regulation of organelles and nuclear condensates^34^. LncRNAs are classified by their genomic locations. *LINC01427* and *LINC00261* are long intergenic ncRNAs (lincRNAs), which are located between protein-coding genes^35^.

It has been demonstrated that lncRNA dysregulation occurs in both animal and human pancreatic islets. Numerous long noncoding RNAs (lncRNAs) are connected to the development of insulin resistance (IR) and are engaged in multiple stages of the insulin manufacturing process^36, 37^. Dysregulated levels of certain lncRNAs in peripheral blood were positively connected with IR, impaired glucose management, inflammation, and transcriptional markers of senescence in a study comparing type 2 diabetes (T2D) patients with healthy controls. Even after controlling for confounding variables, there was a substantial correlation established between this dysregulation and type 2 diabetes^38^. A study by Junpei et al. revealed that a dysregulated lncRNA expression profile is associated with beta cell dysfunction in humans^39^. Beta cell dysfunction leads to impaired insulin secretion, while diminished responsiveness of target tissues to normally secreted insulin leads to insulin resistance. The failure of beta cell function exacerbates insulin resistance, thereby accelerating the progression of type 2 diabetes^40, 33^.

Research on mice has revealed that a specific long non-coding RNA (lncRNA), called lncSHGL, suppresses gluconeogenesis and lipogenesis in the liver^39^. When lncSHGL is restored in mice, it improves hyperglycemia, insulin resistance, and fat accumulation in obese diabetic mice. Conversely, inhibiting lncSHGL in the liver leads to increased fasting blood sugar and fat buildup in normal mice. These results imply that long non-coding RNAs regulate inflammation and the synthesis of fat, which are critical factors in the initiation and development of insulin resistance and glucose regulation^41^.

### SENP2

SENP2 belongs to the family of proteolytic enzymes SUMO-specific proteases (*SENPs*) that reverse the effects of sumoylation, which is a post-translational modification that controls the activity and viability of proteins^42^. Koo YD et al., investigated the role of *SENPs* in energy metabolism. It was found that when a specific muscle cell line was treated with saturated fatty acids, it led to the increase of *SENP2* expression, thereby activating the fatty acid oxidation. Overexpression of *SENP2* in skeletal muscle increased high-fat diet-induced obesity and IR, suggesting a potential therapeutic target for IR-linked metabolic disorders^42^. The study carried out by Chung SS et al reveals that *SENP2* is crucial in adipogenesis regulation. Its expression increases upon adipocyte differentiation and is dependent on protein kinase A activation^43^. Knockdown of *SENP2* in mice leads to the reduction of adipogenesis, PPARgamma, and C/EBPalpha mRNA levels. Sumoylation of C/EBPbeta reverses this effect. Overexpression of C/EBPbeta overcomes knockdown’s inhibitory effect on adipogenesis^43^. Krapf SA et al., studied how (*SENP2*) can influence fatty acid and glucose metabolism in primary adipocytes. The researchers found that silencing of *SENP2* reduced glucose uptake, oxidation and lipogenesis, while increasing lipid oxidation.This suggests that *SENP2* plays a crucial control on energy metabolism in primary human adipocytes^44^.

### SLC22A11

Studies suggest that elevated uric acid levels can lead to metabolic syndrome and insulin resistance, with fructose raising uric acid levels^45^. The protein-coding gene *SLC22A11*, involved in transporting and excreting uric acid, is linked to nephrolithiasis, uric acid, and gout. Its genes *OAT4* and *URAT1* encode renal urate transporters, significantly affecting Serum Urate^46^. Insulin regulates cellular processes through various signaling pathways, with serum- and glucocorticoid-inducible kinases (sgk) acting as mediators^47^. Sgk increases transport activity and OAT4 expression by overcoming Nedd4–2 inhibitory effects^48^. Through the action of ubiquitin ligase Nedd4–2, insulin increases OAT4 expression and transport activity. Nedd4–2 specific siRNA is used to knock down Nedd4–2, weakening regulation. Because insulin competes with sgk2 rather than working through sgk2 to regulate OAT4, the effects of insulin and sgk2 are cumulative^47^. *IGF-1*, a liver hormone, is crucial for metabolism, growth, and development. It increases *OAT4* transport activity and SUMOylation in kidney-derived cells, but PKB-specific inhibitors block its regulation^49^.

The investigation of optimal inheritance models reveals varying modes of genetic inheritance contributing to IR susceptibility. While the recessive model is suitable for rs920590 and rs2078267, the dominant model is favoured for rs7274134 and rs6762208. These findings highlight the complexity of genetic factors influencing IR and the need to consider diverse genetic inheritance patterns in understanding the genetic architecture of metabolic disorders.

This study also emphasizes the significance of clinical parameters in understanding IR development. Serum LDL cholesterol, serum triglycerides, and fat mass exhibit significant associations with IR, indicative of the multifaceted nature of metabolic dysfunction. Combining genomic and clinical data highlights the interaction between hereditary and environmental factors in metabolic health and offers a thorough framework for understanding the pathophysiology of IR^50^.

The validation of genetic markers associated with IR offers insights for personalized therapeutic strategies. By understanding the genetic determinants underlying IR, clinicians can tailor treatment approaches to individual genetic profiles, enhancing the management of metabolic disorders. Identification of novel genetic targets opens avenues for targeted intervention and prevention strategies, paving the way for precision medicine in metabolic health.

While this study provides significant insights into IR’s genetic underpinnings, further investigations are warranted to unravel the intricate interplay between genetic and environmental factors contributing to metabolic dysfunction. Longitudinal studies incorporating larger cohorts and diverse populations are needed to validate and extend these findings. Functional studies are required to understand the molecular mechanisms underlying the observed associations and can provide deeper insights into IR’s pathogenesis. Integrating multi-omics approaches offers a comprehensive understanding of the complex interactions driving metabolic health and disease, facilitating the development of targeted therapeutic interventions and precision medicine strategies aimed at mitigating the burden of metabolic disorders.

## Conclusion

Based on the comprehensive investigation conducted in this study, several key findings have emerged, shedding light on the genetic underpinnings of Insulin Resistance (IR) in the Indian population. Through the identification of the specific SNPs and their associations with IR risk from the literature, this research validated the SNPs role and their complex role in the underlying mechanisms of metabolic disorders. The analysis revealed significant associations between four selected SNPs (rs920590 in *INTS10*, rs7274134 in *LINC01427 - LINC00261*, rs6762208 in *SENP2*, and rs2078267 in *SLC22A11*) and IR status. Notably, the T allele for rs920590, rs7274134, and rs2078267, as well as the C allele for rs6762208, have been associated with an elevated risk of IR in the Indian population.

Furthermore, the determination of optimal inheritance models for each SNP provided additional insights into their genetic inheritance patterns. For instance, the recessive model was identified as the most suitable for rs920590 and rs2078267, while the dominant model was favoured for rs7274134 and rs6762208. These findings suggest varying modes of genetic inheritance contributing to IR susceptibility, highlighting the complexity of genetic factors involved in metabolic dysregulation. The study also highlighted how important it is to consider genetic and clinical factors in determining the etiology of IR and related metabolic disorders. Factors such as serum LDL cholesterol, serum triglycerides, and fat mass exhibited significant associations with IR, emphasising the multifaceted nature of metabolic dysfunction. This study validates the role of specific genetic markers in predisposing individuals to IR in the Indian population, offering valuable insights into the pathogenesis of metabolic disorders. By elucidating the genetic determinants underlying IR, this research lays the foundation for personalised therapeutic strategies tailored to individual genetic profiles, ultimately advancing the management and treatment of metabolic disorders in clinical practice. Further investigations are needed to unravel the intricate interplay between genetic and environmental factors contributing to IR and explore novel avenues for targeted intervention and prevention strategies.

## Data Availability

All data produced in the present study are available upon reasonable request to the authors.

https://www.nugenomics.in/

## Acknowledgements

We would like to express our gratitude to Answer Genomics for supporting us with the research work.

## Abbreviations

(IR): Insulin resistance
(SNPs): single nucleotide polymorphisms
(HOMA2-IR): Homeostasis Model Assessment for Insulin Resistance
(GSA): Global Screening Array
(DM): Diabetes Mellitus
(NCDs): noncommunicable diseases
(BMI): Body mass index
(BF%): body fat percentage
(TG): triglycerides
(LDL-C): high-density lipoprotein cholesterol
(LDL-C): low-density lipoprotein cholesterol
(TC): total cholesterol
(FPG): and fasting plasma glucose
(ORs): odds ratios
(CIs): confidence intervals
(LD): linkage disequilibrium
(HWE): Hardy-Weinberg equilibrium
(AIC): Akaike Information Criterion
(BIC): Bayesian Information Criterion
(T2D): type 2 diabetes

